# Acute Metabolic Emergencies in Diabetes and COVID-19: a systematic review and meta-analysis of case reports

**DOI:** 10.1101/2021.01.10.21249550

**Authors:** Vasileios Papadopoulos, Marios-Vasileios Koutroulos, Dimitra-Georgia Zikoudi, Stefania-Aspasia Bakola, Peny Avramidou, Ntilara Touzlatzi, Dimitrios K. Filippou

## Abstract

**Background:** COVID-19 is associated with DKA (Diabetic Ketoacidosis), HHS (Hyperglycaemic Hyperosmolar State) and EDKA (Euglycaemic DKA). High mortality has been observed in COVID-19-related diabetic ketoacidosis; however, evidence is scarce.

**Methods:** A systematic literature review was conducted using EMBASE, PubMed/Medline, and Google Scholar from January to December 2020 to identify all case reports describing DKA, HHS, and EDKA, in COVID-19 patients. The Joanna Briggs Institute critical appraisal checklist for case reports was used for quality assessment. Univariate and multivariate analysis assessed correlations of study origin, combined DKA/HHS, age, BMI, HbA1c, administered antidiabetics, comorbidities, symptoms onset, disease status (DS), CRP, ferritin, d-dimers, glucose, osmolarity, pH, bicarbonates, ketones, lactates, β-hydroxybutyric acid, anion gap, and acute kidney injury (AKI) with outcome. The relevant protocol was submitted to PROSPERO database (ID: 229356).

**Results:** From 312 identified publications, 41 including 71 cases analyzed qualitatively and quantitatively. The types of acute metabolic emergencies observed were DKA (45/71, 63.4%), EDKA (6/71, 8.5%), combined DKA/HHS (19/71, 26.8%), and HHS (1/71, 1.4%). Overall mortality was 32.4% (22/68 patients; 3 missing). Multivariate analysis by classical regression demonstrated that COVID-19 DS4 (P=3•10^−8^), presence of DKA/HHS (P=0.021), and development of AKI (P=0.037) were all independently correlated with death. Increased DS (P=0.003), elevated lactates (P<0.001), augmented anion gap (P<0.001), and presence of AKI (P=0.002) were associated with DKA/HHS. SGLT-2i administration was linked with EDKA (P=0.004); however, a negative association with AKI was noted (P=0.023).

**Conclusion:** COVID-19 intertwines with acute metabolic emergencies in diabetes leading to increased mortality. Key determinants are critical COVID-19 illness, coexistence of DKA/HHS and AKI. Awareness of clinicians to timely assess them might enable early detection and immediate treatment commencing. As previous treatment with was negatively associated with AKI, thus implying a prophylactic effect on renal function, the issue of discontinuation of SGLT-2i in COVID-19 patients remains to be further evaluated.

**Key messages:** *What is already known on this subject:*

▸ Diabetes mellitus (DM) is a risk factor for poor outcomes in COVID-19 patients.
▸ Diabetic ketoacidosis (DKA) and hyperglycaemic hyperosmolar state (HHS) are not rare in COVID-19 diabetic and non-diabetic patients; key determinants of outcome remain unknown.

*What this study adds:*

▸ COVID-19 intertwines with acute metabolic emergencies in diabetes leading to increased mortality; key determinants are critical COVID-19 illness, coexistence of DKA and HHS as well as development of acute kidney injury.
▸ SGLT2-i administration is linked with euglycaemic DKA in patients with COVID-19, though preserving renal function.

## INTRODUCTION

Diabetes mellitus (DM), especially type 2 diabetes mellitus (T2D), has been identified as a risk factor for poor outcomes in patients with COVID-19 caused by severe acute respiratory syndrome coronavirus 2 (SARS-CoV-2) [1,2]. COVID-19 might either induce new onset diabetes or unmask previously undiagnosed diabetes [3]. Diabetes patients have an increased risk of infection and acute respiratory distress syndrome compared with the general population and the risk is similar [4] or even greater in those with type 1 diabetes mellitus (T1D) than in T2D [5-8]; direct cytopathic effects of SARS-CoV-2 on pancreatic b-cell populations [9] as well as the over-activity of immune system might further explain COVID-19-related severe and resistant to conventional therapy DKA episodes [10]. However, whether SARS-CoV-2 directly infects b-cells in vivo through ACE2 and TMPRSS2 has been debated [11].

COVID-19 is associated with hyperglycaemic emergencies with overrepresentation of T2D in patients presenting with DKA and long-lasting ketosis [12,13]. DKA was the most common reason for hospitalization of T1D patients with COVID-19 [6,14]. Emergency admissions due to acute metabolic crisis in patients with diabetes remain some of the most common and challenging conditions; along with DKA, Hyperglycaemic Hyperosmolar State (HHS) and Euglycaemic DKA (EDKA) are life-threatening different entities. DKA and HHS have distinctly different pathophysiology though sharing basic management protocols. EDKA resembles DKA but without hyperglycaemia [15]. Ketoacidosis is the hallmark of DKA and is attributed to absolute insulin deficiency; therefore, it is found mostly in T1D. DKA is less commonly seen in T2D with triggers such as severe infection [16] and sodium-glucose co-transporter-2 inhibitor (SGLT-2i) therapy [17]. On the other hand, HHS is developed under insulin sufficiency, at least enough to prevent ketosis [15]. As T2D patients tend to be younger, while T1D intertwines with T2D over time in a considerable proportion leading to double or hybrid diabetes [18], DKA is no more specific for T1D, nor HHS for T2D, as the mixed entity (HHS/DKA) is not uncommon.

Predominant features of DKA and EDKA are ketonemia and high anion gap metabolic acidosis. Both DKA and EDKA are defined as pH <7.3 and/or bicarbonate <15 mmol/L, and detection of ketones in blood (ketonemia >3.0 mmol/L) or urine (2+ in urine); however, blood glucose >11 mmol/L is indicative of DKA while <11 mmol/L of EDKA. In contrast, HHS is characterized by very high glucose levels (>33.3 mmol/L) along with very high serum osmolality (> 320 mOsm/kg). Several case reports concerning acute emergencies related to glucose metabolism in COVID-19 patients have been reported. Additionally, high mortality in COVID-19 and DKA has been reported [19]. A systematic review concluded that mortality rate from DKA among COVID-19 patients might approach 50% and insisted on differentiating isolated DKA from combined DKA/HHS as the latter, which represents nearly one-fifth of the DKA cases, tends to have higher mortality than DKA alone [20]. Diabetic COVID-19 patients should be assessed for disease severity and presence of complications of diabetes, while undiagnosed diabetes should be considered, especially in patients feeling unwell [21].

The present systematic review and meta-analysis aimed to identify all case reports describing DKA, EDKA, HHS, and DKA/HHS, in patients with confirmed COVID-19 infection and provide further evidence by describing both primary (survival/discharge vs. death) and secondary (type of metabolic emergency) outcome in relation with origin, coexistence of DKA/HHS, age, body mass index (BMI), HbA1c, prior administration of antidiabetic treatment, comorbidities, days from onset of symptoms, disease status (DS), C-reactive protein (CRP), ferritin, d-dimers, glucose, osmolarity, arterial pH, bicarbonates, ketones, lactates, β-hydroxybutyric acid (β-HB), anion gap, as well as acute kidney injury (AKI).

## METHODS

### Literature search

A systematic literature review was conducted using EMBASE and PubMed/Medline from January 2020 to December 2020 to identify all case reports describing DKA, EDKA, HHS, and DKA/HHS, in patients with confirmed COVID-19 infection through positive RT-PCR for SARS-CoV-2 RNA in nasopharyngeal swab or bronchoalveolar lavage, using the search strategy that included the terms (diabetes AND ketoacidosis AND covid) OR (diabetic AND ketoacidosis AND covid) OR (euglycemic AND diabetic AND ketoacidosis AND covid) OR (hyperglycaemic AND hyperosmolar AND state AND covid). Google Scholar database was used as an additional pool of published data; iterative search was performed until no additional publication could be traced. Personal communication was attempted where necessary. Unpublished dissertations and other unpublished work were scavenged. No software was used for study retrieval. The review methods were established prior to the conduct of the review. No significant deviations from the protocol were allowed. No funding was received. The relevant protocol was submitted to PROSPERO database on January 5, 2021 and corrected on January 8, 2021 (ID: 229356).

### Study selection

Eligible studies were all that (1) described one or more case reports containing requested data retrievable at individual level; (2) were written in English, (3) had a consistent outcome of interest and (4) were not duplicates of older versions. No restrictions were considered regarding publication time as no outdated studies existed due to the novelty of the topic. Due to the inflated interest on publishing new works on the field, we proceeded to pre-run searches prior to the final analysis aiming to include any further studies identified. No software was used for recording decisions; all data were transformed to a Word table. Sources of financial support were traced where possible.

### Outcome measures

The present study was conducted in accordance to the PRISMA statement for systematic reviews and meta-analyses [22]. Both primary (survival/discharge vs. death) and secondary (type of metabolic emergency) outcome was assessed in relation with origin, coexistence of ketotic and hyperosmotic state, age, BMI, HbA1c, prior administration of antidiabetic treatment including insulin, metformin, sulfonylureas, dipeptidyl peptidase-4 inhibitors (DPP-4i), glucagon-like peptide-1 receptor agonists (GLP-1 RAs), SGLT-2i, and pioglitazone, comorbidities including T1D/T2D, arterial hypertension, hyperlipidemia, coronary artery disease, asthma and others, days from onset of symptoms, DS (as described in the Supplemental Material), CRP, ferritin, d-dimers, glucose, osmolarity, arterial pH, bicarbonates, ketones, lactates, β-HB, anion gap, as well as AKI. AMSTAR 2 checklist was used to assess the quality of the present study [23,24].

### Data extraction

A structured data collection form was used to extract all necessary data from each study: study title, first author, DKA/HHS presence, age, BMI, HbA1c, antidiabetic treatment, comorbidities, days from onset of symptoms, DS, CRP, ferritin, d-dimers, glucose, osmolarity, arterial pH, bicarbonates, ketones, lactates, β-HB, anion gap, AKI, and outcome. AMSTAR 2 checklist was used to assess quality of the present study

### Quality assessment of the studies

The Joanna Briggs Institute (JBI) critical appraisal checklist for case reports, which addresses internal validity and risk of bias of case reports designs, particularly confounding and information bias, in addition to the importance of clear reporting, was used for quality assessment [26-29]. All studies that failed to fulfill requirements of first six questions were considered as of “suboptimal quality”; controversially, an “optimal quality” remark was given.

### Pattern of collaboration

Six reviewers (V.P., M.-V.K., D.-G.Z., S.-A.B., P.A., and N.T.) performed study selection, data extraction, and quality assessment working simultaneously as three independent couples (one for screening and the other for checking decisions). These couples were blinded to each other’s decisions. D.F. was responsible to dissolve any disagreement.

### Data synthesis and statistical analysis

Classical regression without weighing each data point was used for data synthesis. The relevant odds ratios (OR) were used to construct a forest plot for visualization purposes using Revman 5.3 software [30]. Further details are provided in as Supplemental Material.

### Patient and Public Involvement Statement

Patients and the public were not involved in any way in the present study.

## RESULTS

### Study characteristics

During the final pre-run search prior to the final analysis carried out on January 7, 2021, 312 potentially relevant publications were identified through a thorough search of literature; 174 in EMBASE, 138 in PubMed/Medline, while two more publications of interest were indentified through Google Scholar. No unpublished data of interest was detected. Personal contact contributed further information. Fourty-one publications referring to 71 separate case reports included in qualitative and quantitative synthesis [3, 31-70] (Figure 1). Three case reports did not refer to COVID-19 patients (Supplemental Table 1). All study characteristics are analytically presented in Table 1. The quality of the present study was evaluated as “high” using the AMSTAR 2 checklist.

**Table 1.**
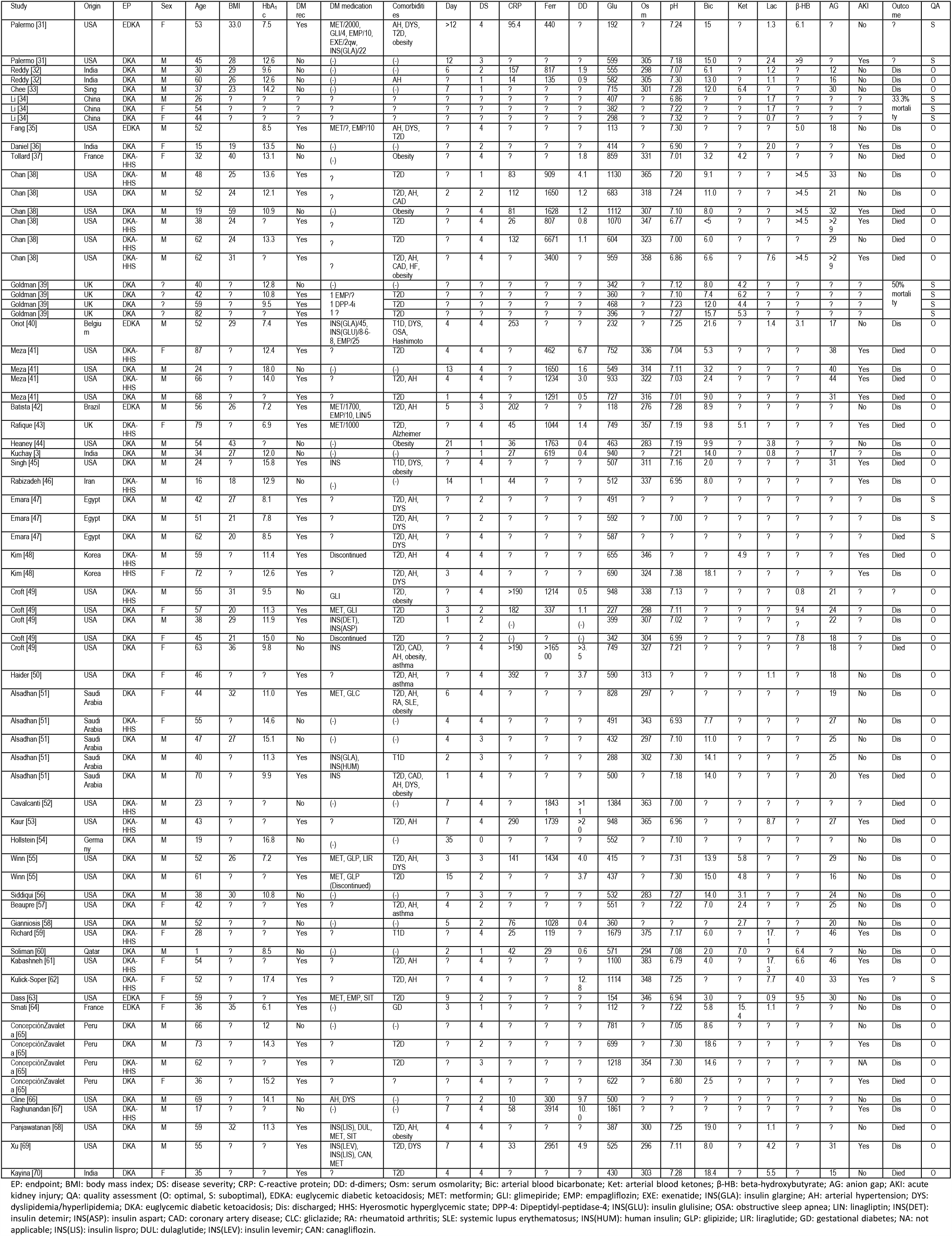
Studies included in the present meta-analysis along with analytical description of patients characteristics.

**Figure 1.**
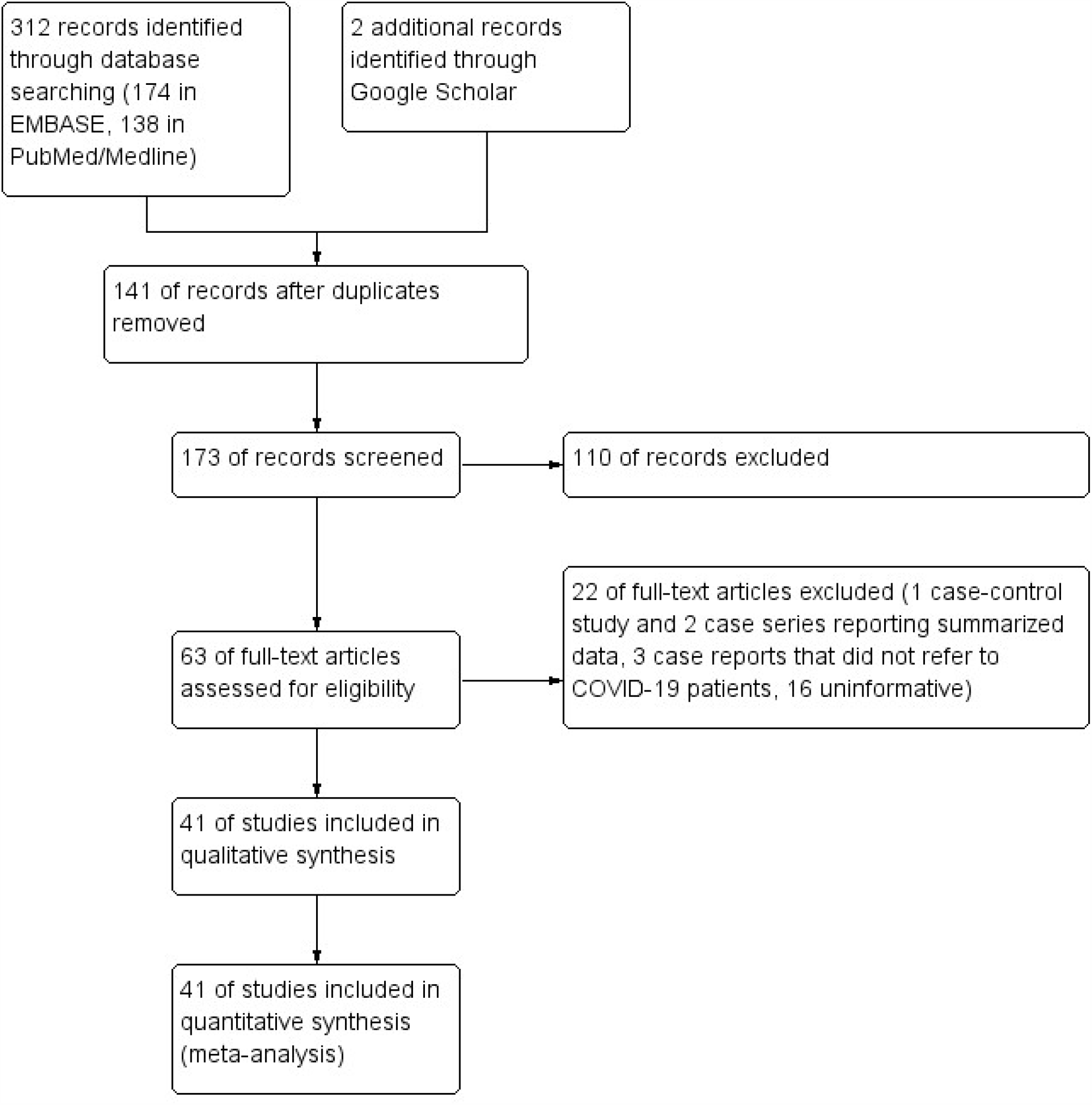
Study flow diagram

### Quality assessment of the studies

Quality remarks are provided in Table 1; all details concerning quality assessment items are depicted analytically in Table 2. The inter-rater agreement between the two authors carried out the quality assessment process was high, as kappa was 0.87 (95% CI: 0.79-0.95). There was no difference between studies of “optimal” and “suboptimal” quality regarding outcome (P=0.756).

**Table 2.** Quality assessment items of JBI critical appraisal list for case reports concerning included studies.

| Study | Were patient's demographic characteristics clearly described? | Was the patient's history clearly described and presented as a timeline? | Was the current clinical condition of the patient on presentation clearly described? | Were diagnostic tests or assessment methods and the results clearly described? | Was the intervention(s) or treatment procedure(s) clearly described? | Was the post-intervention clinical condition clearly described? | Were adverse events (harms) or unanticipated events identified and described? | Does the case report provide takeaway lessons? | Optimal quality |
| --- | --- | --- | --- | --- | --- | --- | --- | --- | --- |
| Palermo | Yes | U | Yes | Yes | U | No | U | Yes | No |
| Reddy | Yes | Yes | Yes | Yes | Yes | Yes | No | Yes | Yes |
| Chee | Yes | Yes | Yes | Yes | Yes | U | No | Yes | Yes |
| Li | Yes | Yes | Yes | Yes | No | No | No | Yes | No |
| Fang | Yes | Yes | Yes | Yes | Yes | Yes | Yes | Yes | Yes |
| Daniel | Yes | U | Yes | Yes | Yes | Yes | No | Yes | Yes |
| Tollard | Yes | Yes | Yes | Yes | Yes | Yes | No | Yes | Yes |
| Chan | Yes | Yes | Yes | Yes | Yes | Yes | No | Yes | Yes |
| Goldman | No | Yes | Yes | Yes | U | No | No | Yes | No |
| Oriot | Yes | Yes | Yes | Yes | Yes | Yes | U | Yes | Yes |
| Meza | Yes | Yes | Yes | Yes | Yes | Yes | No | Yes | Yes |
| Battista | Yes | Yes | Yes | Yes | Yes | Yes | U | Yes | Yes |
| Rafique | Yes | Yes | Yes | Yes | Yes | Yes | No | Yes | Yes |
| Heaney | Yes | Yes | Yes | Yes | Yes | Yes | No | Yes | Yes |
| Kuchay | Yes | Yes | Yes | Yes | Yes | Yes | No | Yes | Yes |
| Singh | Yes | Yes | Yes | Yes | Yes | Yes | U | Yes | Yes |
| Rabizadeh | Yes | Yes | Yes | Yes | Yes | Yes | No | Yes | Yes |
| Emara | Yes | Yes | Yes | Yes | No | Yes | No | Yes | No |
| Kim | Yes | Yes | Yes | Yes | Yes | Yes | No | Yes | Yes |
| Croft | Yes | Yes | Yes | Yes | U | U | No | Yes | Yes |
| Haider | Yes | Yes | Yes | Yes | Yes | Yes | No | Yes | Yes |
| Alsadhan | Yes | Yes | Yes | Yes | Yes | Yes | No | Yes | Yes |
| Cavalcanti | Yes | Yes | Yes | Yes | U | Yes | No | Yes | Yes |
| Kaur | Yes | Yes | Yes | Yes | Yes | Yes | No | Yes | Yes |
| Hollstein | Yes | Yes | Yes | Yes | Yes | Yes | No | Yes | Yes |
| Winn | Yes | Yes | Yes | Yes | Yes | Yes | No | Yes | Yes |
| Siddiqui | Yes | Yes | Yes | Yes | Yes | Yes | No | Yes | Yes |
| Beaupre | Yes | Yes | Yes | Yes | Yes | Yes | No | Yes | Yes |
| Gianniosis | Yes | Yes | Yes | Yes | Yes | Yes | No | Yes | Yes |
| Richard | Yes | Yes | Yes | Yes | Yes | Yes | No | Yes | Yes |
| Soliman | Yes | Yes | Yes | Yes | Yes | Yes | No | Yes | Yes |
| Kabashneh | Yes | Yes | Yes | Yes | Yes | Yes | No | Yes | Yes |
| Kulick-Soper | Yes | Yes | Yes | Yes | U | No | No | Yes | No |
| Dass | Yes | Yes | Yes | Yes | Yes | Yes | Yes | Yes | Yes |
| Smati | Yes | Yes | Yes | Yes | Yes | Yes | No | Yes | Yes |
| Zavaleta | Yes | Yes | Yes | Yes | Yes | U | No | Yes | Yes |
| Cline | Yes | Yes | Yes | Yes | Yes | Yes | No | Yes | Yes |
| Raghunandan | Yes | Yes | Yes | Yes | Yes | Yes | Yes | Yes | Yes |
| Panjawatanan | Yes | Yes | Yes | Yes | Yes | Yes | No | Yes | Yes |
| Xu | Yes | Yes | Yes | Yes | Yes | Yes | No | Yes | Yes |
| Kayina | Yes | Yes | Yes | Yes | Yes | Yes | No | Yes | Yes |

Y: Yes; N: No; U: Unclear

### Primary outcome

The types of acute metabolic emergencies observed were DKA (45/71, 63.4%), EDKA (6/71, 8.5%), combined DKA/HHS (19/71, 26.8%), and HHS (1/71, 1.4%). Overall mortality was 32.4% (22/68 patients; 3 missing).

Absence of combined DKA/HHS (P=0.006), and absence of AKI (P=0.001) are correlated with increased OR for survival. Among patients with T1D or T2D, administration of insulin was associated with an increased OR of succumbing (5.99; 95% CI: 0.48-76.9). In contrast, T2D patients who received metformin had an OR of 0.14 (95% CI: 0.01-2.52).

BMI (P=0.071), new onset of diabetes, either T1D or T2D (P=0.071), DS (P=0.083), osmolarity (P=0.076), pH value (P=0.063), and β-HB (P=0.052) were considered needing further evaluation and thus were included in multivariate regression analysis. Detailed univariate analysis concerning correlations of patients characteristics with outcome is analytically presented at Table 3 and Figure 2. The most parsimonious multivariate model is highly significant (P=10^−4^), suggesting that COVID-19 DS4 (P=3•10^−8^), presence of DKA/HHS (P=0.021), and development of AKI (P=0.037) are all independently correlated with death (Figure 3).

**Table 3.**
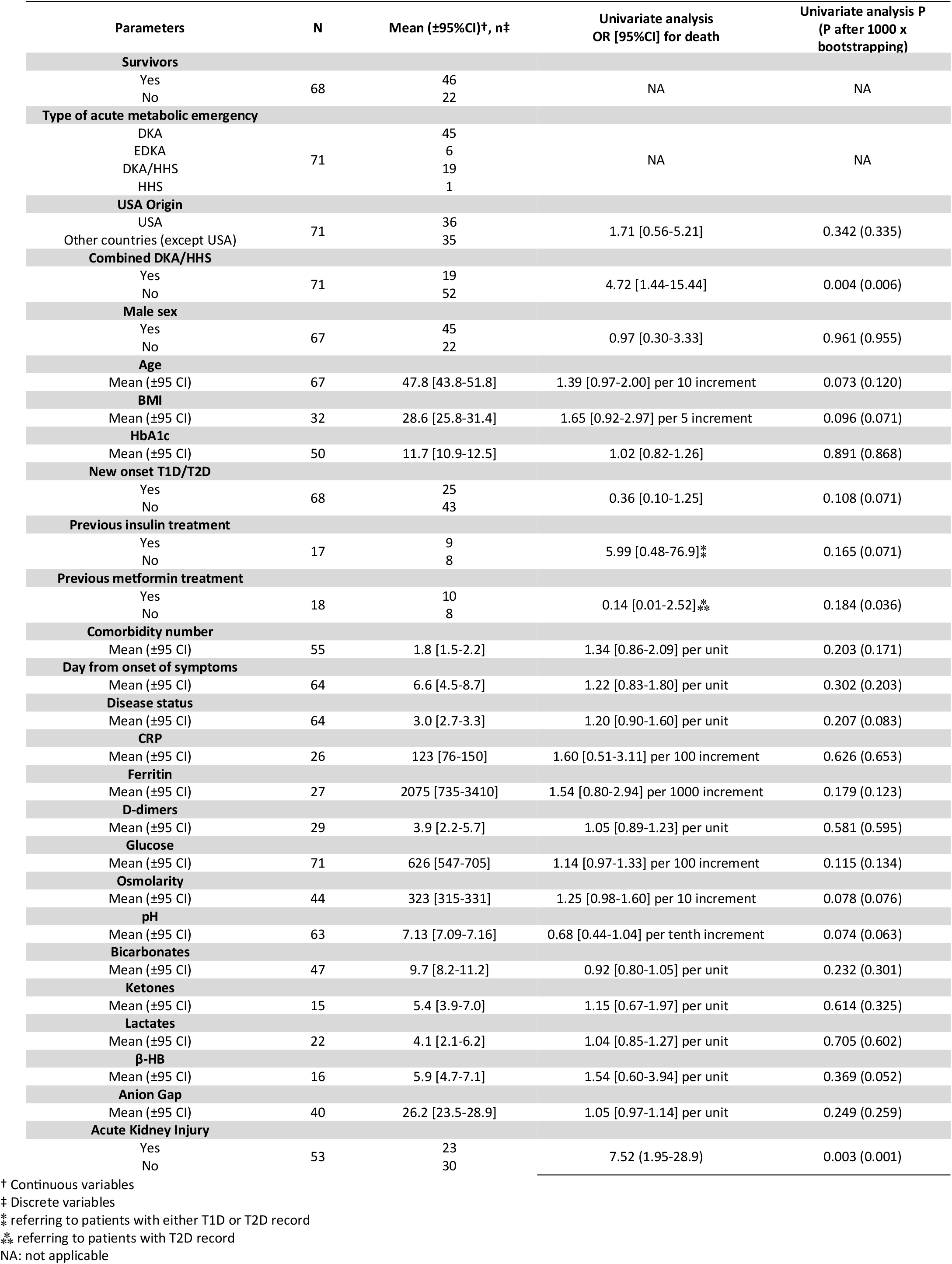
Characteristics of COVID-19 patients in relation with outcome.

**Figure 2.**
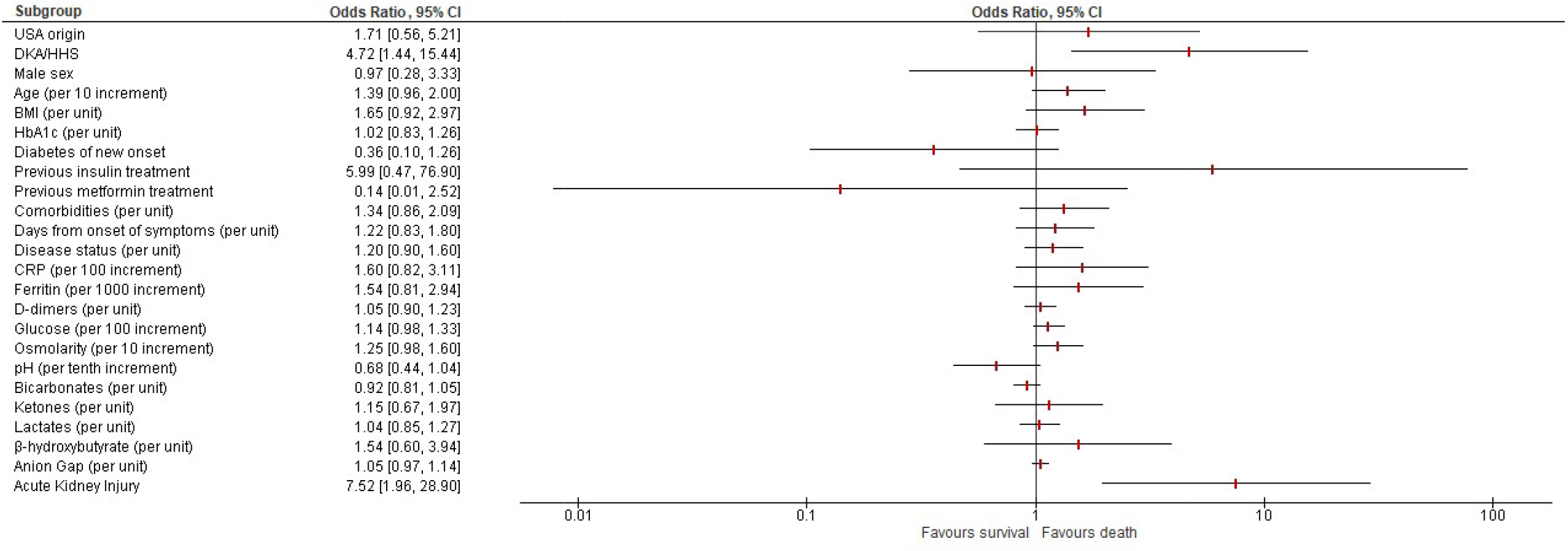
Forest plot depicting various patients’ characteristics odds ratios OR for death.

**Figure 3.**
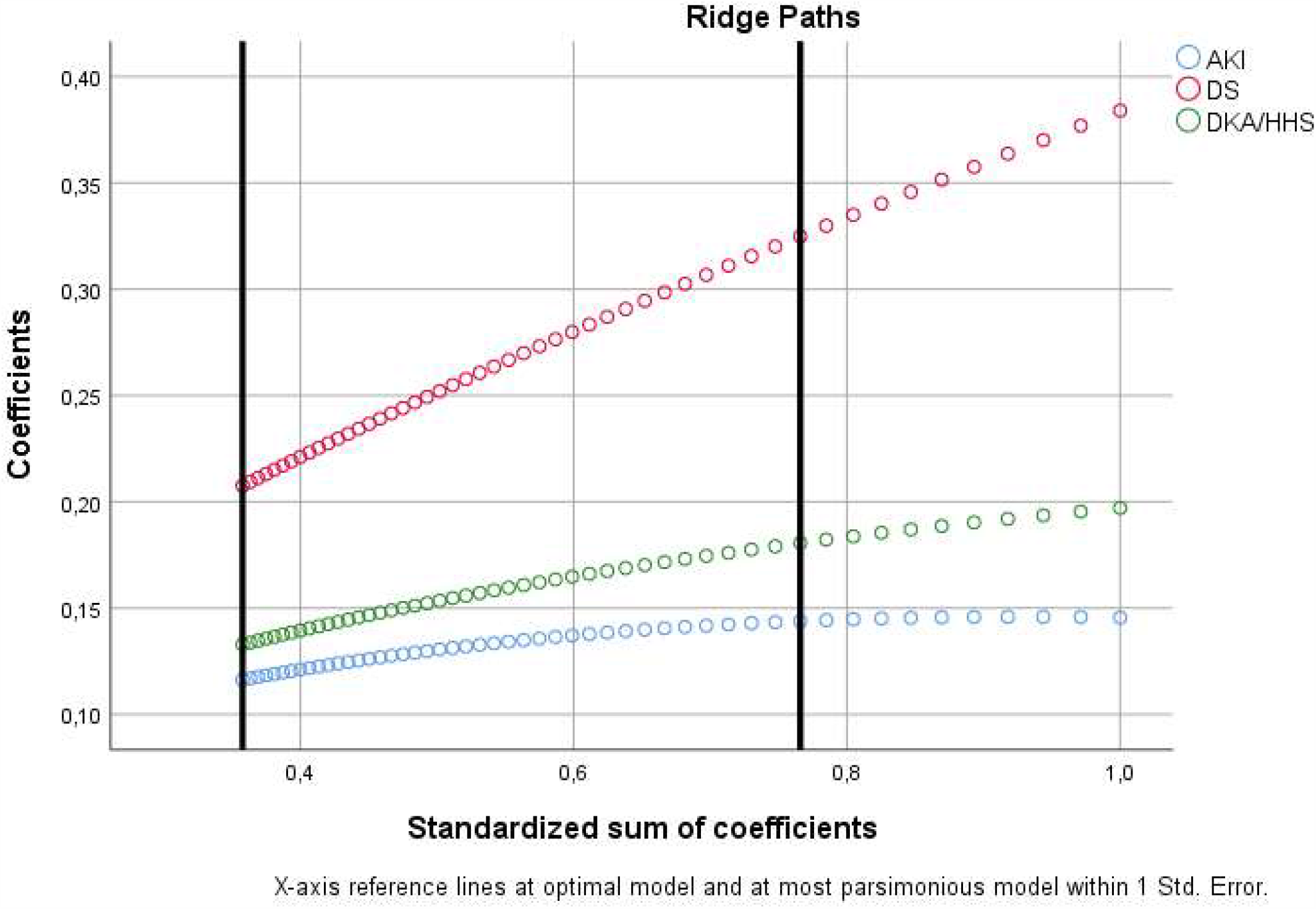
Ridge paths as derived from optimal scaling regularization process; the most parsimonious multivariate model is highly significant (P=10^−4^), suggesting that COVID-19 Disease Severity 4 (P=3•10^−8^, tolerance: 0.838), simultaneous presence of DKA and HHS (P=0.021, tolerance: 0.803), and development of AKI (P=0.037, tolerance: 0.788) are all independently correlated with death.

### Secondary outcome

Increased DS (P=0.003), elevated lactates (P<0.001), augmented anion gap (P<0.001), and presence of AKI (P=0.002) were associated with DKA/HHS.

SGLT-2i administration was linked with EDKA (Fisher’s exact P=0.004); however, a negative association with AKI was noted (P=0.023).

## DISCUSSION

We describe three major determinants of outcome during acute metabolic emergencies in diabetes in COVID-19 patients: i) COVID-19 critical illness necessitating mechanical ventilation (DS4), ii) simultaneous presence of ketoacidosis and hyperosmosis as DKA/HHS (P=0.021), and iii) AKI. To the best of our belief, this is the first time that prominent features that enable intertwining of COVID-19 and acute metabolic emergencies in diabetes leading to increased mortality are presented in a comprehensive and justified manner.

Our study revealed an overall mortality of 32.4% (22/68 patients; 3 missing) in COVID-19 patients who had developed DKA, EDKA, DKA/HHS, and HHS. Interestingly, two recently published case series report mortality rates that range from as low as 7.7% (2/26 patients) [12] and 1/7 patients (12.9%) [71], to 50% (25/50 patients) [19]; however, whether these case series are representative remains to be answered.

Our findings regarding the independent correlation of critical illness and mortality during COVID-19-related acute metabolic emergencies in diabetes confront with what is already reported [19]. Survivors were reported to have lower CRP levels when compared with non-survivors; additionally, the former required intubation and mechanical ventilation more frequently [19].

Furthermore, we exhibited an independent correlation of AKI with mortality during COVID-19-related DKA, EDkA, DKA/HHS, and HHS. AKI is quite common among patients without critical illness and usually has a mixed etiology intertwining sepsis, ischemia and nephrotoxicity and perplexing recognition and treatment [72]. AKI was observed in 92% (23/25) and 60% (15/25) of COVID-19-related DKA non-survivors and survivors, respectively; these data indicate that AKI is significantly correlated with death in patients with COVID-19-related DKA [19]. Similarly, renal replacement therapy was required in 40% (10/25) and 4% (1/25) COVID-19-related DKA non-survivors and survivors, respectively, implying that renal replacement therapy is significantly correlated with death in these patients [19].

Additionally, we demonstrated an independent correlation of mixed DKA/HHS related to COVID-19 infection with non-surviving. Our results are in keeping with Pal et al., who describe a statistically significant difference in arterial blood pH between COVID-19-related DKA survivors when compared with non-survivors [20]. Despite that data linking increased osmolarity with increased fatality rate in COVID-19 patients are lacking, it is well known that death occurs in 5–16% of patients with HHS in general, a rate that is about 10-fold higher than that reported for DKA [73-75]. Moreover, a hypertonic environment impairs the immune response, thus facilitating the development of infection [76].

We have noticed that all patients but one (a patient with gestational diabetes) who had developed EDKA received SGLT-2i treatment; however, a negative association with AKI was noted thus implying a prophylactic effect on renal function. Treatment with SGLT-2i has been reported to trigger EDKA in T2D patients with [39] or without COVID-19 infection, usually during other infections, sepsis or surgery [77-79]. These regimens might be prescribed even for T1D, even without reimbursement; interestingly, a single case report of a patient with T1D who received empagliflozin 25mg q24h and developed EDKA during COVID-19 pneumonia has been recently published [40]. Glycaemic stability can mislead the clinician, since hyperglycosuria induced by SGLT-2i may blunt hyperglycaemia during infection and contribute to a lack of insulin, finally promoting ketogenesis [77]. Therefore, it is strongly advised that the use of SGLT-2i should be discontinued at once as soon as COVID-19 is diagnosed, while the exclusive administration of insulin is considered the safest choice [79-80]. Nevertheless, our finding deserves further evaluation being in keeping with the fact that SGLT2-i administration slows the decline observed in the annual renal function in T2D patients with eGFR<60 ml/min/1.73m^2^, in non-COVID-19 patients [81].

We have not detected any correlation of antidiabetic drug category other than SGLT-2i with any special type of metabolic emergency or outcome; this observation conveys the limitations of the small sample size of the present study. GLP-1 RAs reduce circulating inflammatory biomarkers in diabetic and/or obese patients while insulin reduces these biomarkers in critically ill patients. Pioglitazone was also shown to upregulate ACE2 in hepatocytes of rats fed with a high fat diet. Finally, the DPP-4 is the entry receptor of MERSCoV, raising concerns about the impact of DPP-4i during the course of coronavirus infection [82].

Interestingly, we demonstrated that insulin-treated patients presented an increased OR to succumb in contrast with those treated with metformin. Though insulin administration had been associated with poor prognosis by another group of investigators [83] it restores ACE and ACE2 serum levels, thus hypothesizing that it exerts a protective effect at least in patients that are non-insulin-depleted [84]. Therefore, this finding of ours could reflect a confounder effect due to either the type of diabetes, or increased age. T1D patients, who are by definition insulin-dependent, when compared with T2D patients, are more prone to adverse outcome during COVID-19 infection [5]; moreover, unfavorable outcome was observed more often in older patients presenting COVID-19-related DKA [19].

COVID-19 might either induce new onset diabetes or unmask previously undiagnosed T1D or T2D; elevated HbA1c values at admission confronts for the latter [3]. Both SARS and COVID-19 have been reported to trigger transient insulin resistance and hyperglycaemia. SARS results in elevated glucose during admission; however, glucose intolerance is resolved at hospital discharge [39]. SARS-CoV-2 can trigger severe diabetic ketoacidosis at presentation in individuals with new-onset diabetes despite that evidence etiologically linking SARS-CoV-2 with T1D are lacking [85]. This COVID-19 induced insulin resistance may partly explain poor responses to DKA management [39]. Emerging data indicate a bidirectional relationship between T2D and COVID-19 [86]. Impairment of innate and adaptive immunity tames the ability to fight infection in patients with diabetes and particularly in obese. Furthermore, severe COVID-19 infection significantly reduces the numbers of natural killer cells (CD4+/CD8+ cells) and CD4+/CD8+ lymphocytes. The association between COVID-19 and hyperglycemia in elderly patients with T2D might reflect metabolic inflammation and exaggerated cytokine release. SARS-CoV2 infection can deteriorate glycemic control by enhancing insulin resistance and impaired insulin secretion, thus leading to DKA [10,87]. The unique interactions between SARS-CoV-2 and the RAAS might provide yet another mechanism in the pathophysiology of DKA firstly by direct entry of SARS-CoV-2 into pancreatic islet cells worsening b-cell injury and secondly by downregulation of ACE2 after viral entry that can lead to unopposed angiotensin II and subsequent insulin secretion impedance [88]. ACE2 expression at both mRNA and protein level is increased substantially in human b-cells in response to inflammatory cytokines, presumably rendering these cells more susceptible to infection [89].

Nevertheless, rigorous and proper restoration of volume and insulin adequacy, along with potassium preservation should be commenced immediately in any case of COVID-19-related DKA. As the relationship between SARS-CoV-2 and the RAAS can complicate DKA management due to increased pulmonary vascular permeability and worsened damage to lung parenchyma, fluid replacement needs to be administered judiciously to avoid aggravating pulmonary injury. This also underlies the importance of careful assessment of fluid status through objective hemodynamic parameters to determine the amount of fluid replacement. Another important aspect in DKA management is monitoring and correcting electrolyte abnormalities. As angiotensin II stimulates aldosterone secretion and increases renal potassium loss, hypokalemia might evolve necessitating additional potassium supplementation in order to continue intravenous insulin to suppress ketogenesis [86,90-92].

A major limitation of the present study is that it relies only in case reports, which lack the ability to generalize or establish cause-effect relationship, while conveying all danger of over-interpretation, publication bias, retrospective design, and distraction of reader when focusing on the unusual. However, the major merits of case reporting focus, among other, on detecting novelties, and generating hypotheses, which are considered absolutely necessary during the course of COVID-19 pandemic [93].

As a conclusion, COVID-19 intertwines with acute metabolic emergencies in diabetes leading to increased mortality. Key determinants are critical COVID-19 illness, coexistence of ketoacidosis and hyperosmosis and AKI and awareness of clinicians to timely assess them might enable early detection and immediate treatment commencing of DKA, EDKA, HHS and DKA/HHS in COVID-19 patients. Moreover, as previous treatment with SGLT-2i demonstrated a negative association with AKI thus implying a prophylactic effect on renal function the issue of discontinuation of these regimens in COVID-19 patients remains to be further evaluated. Properly designed studies are needed to consolidate knowledge on the underlying pathophysiological mechanisms.

## Data Availability

All data referred to the manuscript will be available at https://www.enargeia.eu/.

## Contributors

VP was responsible for study design, data interpretation, as well as statistical analysis and participated in study selection, data extraction, quality assessment, as well as the writing of the manuscript. MVK, DGZ, SAB, PA, NT, participated in study selection, data extraction, quality assessment, and the writing of the manuscript; DF supervised the project and helped develop the idea of the project.

## Patient consent for publication

Not required.

## Provenance and peer review

Not commissioned; externally peer reviewed.

## Data availability statement

Data are available in a public, open access repository. Not applicable.

## SUPPLEMENTAL MATERIAL

### Disease status

The disease-status (DS) of COVID-19 patients was classified based on the adaptation of the Sixth Revised Trial Version of the Novel Coronavirus Pneumonia Diagnosis and Treatment Guidance, as described previously [25]. Mild cases (DS1) were defined as mild clinical symptoms (fever, myalgia, fatigue, diarrhea) and no sign of pneumonia on thoracic X-Ray or/and CT scan. Moderate cases (DS2) were defined as clinical symptoms associated with dyspnea and radiological findings of pneumonia on thoracic X-Ray or/and CT scan, and requiring a maximum of 3 L/min of oxygen. Severe cases (DS3) were defined as respiratory distress requiring more than 3 L/min of oxygen and no other organ failure. Critical cases (DS4) were defined as respiratory failure requiring mechanical ventilation, shock and/or other organ failure that require an intensive care unit (ICU).

### Data synthesis and statistical analysis

As variability values associated to each study were lacking, neither data synthesis nor classical meta-regression was applicable for case reports. Therefore, our option was to use classical regression without weighing each data point; this procedure still offered the advantage of a useful insight in cases that values were not too spread, despite that precise estimations could not be achieved. Thus, univariate analysis was performed to assess potent correlations of independent variables (study origin, presence of combined DKA/HHS, age, BMI, HbA1c, previously administered antidiabetics, comorbidities, days from onset of symptoms, DS, CRP, ferritin, d-dimers, glucose, osmolarity, arterial pH, bicarbonates, ketones, lactates, β-HB, anion gap, AKI) with outcome (survival/discharge or death), which was considered as dependent variable, with the aid of binary regression. At a next step, multivariate analysis was performed with binary logistic regression over discretized, imputed, and regularized data; outcome was considered as a dependent variable, while all variables that reached a level of statistical significance <0.10 in the univariate analysis were treated as potent independent ones (the probability for stepwise entry and removal were set to 0.05 and 0.10 accordingly, the classification cutoff was set to 0.5 and the maximum number of iterations was set to 20). During this process, ridge regression was used to avoid model overfitting, tolerate large variances and overcome collinearity obstacles, all at the least possible additive bias; all variables of interest, if not already binary, were transformed to binary ones through nominal optimal scaling along with discretization to two groups, imputing of missing data was added, and 10-fold crossvalidation was selected through Optimal Scaling procedure (SPSS CATREG option). Models including parameters with tolerance <0.67 (variance inflation factor >1.5), as deduced from corresponding linear regression analysis assessing numerical values to outcome, were rejected to avoid collinearity. Imputed data tolerated missing values at a maximum of 25% for the entire model. Univariate comparisons were performed with the use of Pearson’s χ^2^ for discrete variables; Fisher’s exact test was alternatively preferred in case that expected frequencies were <5 in more than one cell. Kappa statistics were used for the evaluation of inter-rater agreement between authors. The level of statistical significance was set to p=0.05; values of 0.05≤p≤0.1 were considered as needed further evaluation. Means are accompanied by their 95% confidence intervals (CI). All numerical values are given with at least two significant digits. Statistical analysis was performed with the use of IBM SPSS Statistics software, version 26.0.0.0, for Windows (IBM Corp ©) and MedCalc® Statistical Software version 19.6 (MedCalc Software Ltd, Ostend, Belgium; https://www.medcalc.org; 2020). The relevant odds ratios (OR) were used to construct a forest plot for visualization purposes using Revman 5.3 software from the Cochrane Collaboration [30].

### Supplemental Tables

**Supplemental Table 1.**
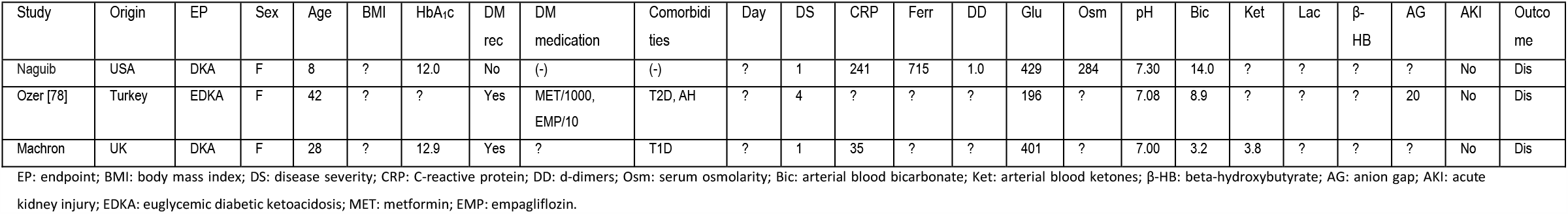
Case report studies excluded in the present meta-analysis (n=3).

